# Trends in using intraoperative parathyroid monitoring during parathyroidectomy: Protocol and rationale for a cross-sectional survey study of North American surgeons

**DOI:** 10.1101/2024.03.13.24304237

**Authors:** Phillip Staibano, Tyler McKechnie, Alex Thabane, Michael Xie, Han Zhang, Michael K. Gupta, Michael Au, Jesse D. Pasternak, Sameer Parpia, JEM (Ted) Young, Mohit Bhandari

## Abstract

**Introduction:** Hyperparathyroidism is a common endocrine disorder that can be secondary to a single or multiple abnormal parathyroid glands and can occur in the context of chronic kidney disease (CKD). There are three types of hyperparathyroidism, and all are definitively managed via surgical extirpation of abnormal parathyroid gland tissue. Intraoperative parathyroid hormone (IOPTH) monitoring was introduced over three decades ago and has been shown to improve clinical outcomes in patients with primary hyperparathyroidism (PHPT). As the incidence of PHPT rises due to improving screening globally and the incidence of CKD rises, it will be important to optimize adoption and standardization of IOPTH within endocrine surgery centers around the world. We will perform a cross-sectional survey study of surgeon rationale, operational details, and barriers associated with IOPTH adoption across North America.

**Methods and analysis:** We will utilize a convenience sampling technique to distribute an online survey to head and neck surgeons and endocrine surgeons across North America. This survey will be distributed via email to three North American professional societies (i.e., Canadian Society for Otolaryngologists–Head and Neck Surgeons, American Head and Neck Society, and American Association of Endocrine Surgeons). The survey will consist of 30 multiple choice questions that are divided into three concepts: (1) participant demographics and training details, (2) details of surgical adjuncts during parathyroidectomy, and (3) barriers to adoption of IOPTH. Descriptive analyses and multiple logistic regression models will be used to evaluate the impact of demographic, institutional, and training variables on the use of IOPTH monitoring and barriers to IOPTH adoption.

**Discussion:** This study will explore IOPTH monitoring for guiding parathyroid surgeries in secondary and tertiary hyperparathyroidism. An ability to capture surgeon practices regarding IOPTH monitoring will inform trials aimed to help optimize IOPTH in challenging populations.

**Ethics and dissemination:** Ethics approval was obtained by the Hamilton Integrated Research Ethics Board (2024-17173-GRA). We do not expect any survey respondents to experience any harms because of participating in this study. We plan to present the results of this study at national and international conferences, and we will publish these findings in peer-reviewed surgical journals. We plan to use these study findings to advocate for adoption of IOPTH technologies and inform future studies and trials.

## INTRODUCTION

Hyperparathyroidism is defined by pathological calcium homeostasis related to abnormal secretion of parathyroid hormone (PTH) [1]. Primary hyperparathyroidism is caused by parathyroid adenomatous disease of a single gland in over 80% of cases. It is classically associated with osteoporosis, nephrocalcinosis, and nephrolithiasis [2]. In contrast, secondary and tertiary hyperparathyroidism are diagnosed in the context of chronic kidney disease (CKD) or following renal transplant, respectively. These diseases are related to hyperplasia from chronic hypercalcemia in the former secondary disease, and autonomously functioning hyperplastic glands in the case of post-transplant tertiary hyperparathyroidism [3]. The global incidence of primary hyperparathyroidism is expected to increase alongside wider accessibility to serum calcium measurements and it remains one of the most common endocrine disorders amongst postmenopausal women [4]. It is well-established that secondary hyperparathyroidism is associated with worse cardiovascular and mortality outcomes in CKD and will likely rise in prevalence alongside increasing rates of CKD; further tertiary disease is becoming more prevalent given greater access to kidney transplant [5, 6]. Primary and tertiary hyperparathyroidism requires surgical exploration and extirpation of one or more parathyroid glands, while in certain cases, secondary hyperparathyroidism can be managed medically [7]. Parathyroidectomy is a commonly performed surgery within the technical scope of otolaryngology–head and neck surgeons and endocrine surgeons [1, 8]. Despite advances in imaging that have improved cure rates in primary hyperparathyroidism, the surgical management of secondary and tertiary hyperparathyroidism remains challenging [9].

In 1991, Irvin and colleagues successfully harnessed the rapid half-life (i.e., 3–5 minutes) of PTH by performing intraoperative PTH (IOPTH) monitoring to guide parathyroidectomy [10]. Since then, this technology has been used increasingly in parathyroidectomy with recent meta-analyses demonstrating its utility in reducing persistent and recurrent disease following surgery [11, 12]. The benefit, however, of IOPTH monitoring in surgery for secondary and tertiary hyperparathyroidism remains controversial [13]. Herein, we report a protocol for cross-sectional survey study of IOPTH monitoring practices amongst head and neck and endocrine surgeons across North America. The aims of this study are multifold: (1) To describe the clinical indications and technical details regarding IOPTH monitoring in primary, secondary, and tertiary hyperparathyroidism; (2) To identify the clinical, geographic, and institutional barriers to adopting IOPTH monitoring at endocrine centres across North America.

## METHODS AND ANALYSIS

### Sampling technique and recruitment

We will perform convenience sampling to recruit survey respondents for this descriptive survey study. All candidate respondents will be head and neck surgeons and endocrine surgeons practicing in North America and they will be contacted via the email lists of professional and academic societies (e.g., American Head and neck Society [AHNS], American Association of Endocrine Surgeons [AAES], and Canadian Society of Otolaryngologists–Head and Neck Surgeons [CSO]). It is important to note that active AAES members may reside in North, Central, and South America [14]. As these professional societies may not delineate staff surgeons from surgical trainees, this will be accounted for with the survey and planned analysis. Our purpose is to sample academic surgeons who work at high-volume endocrine centers and so, distributing this survey via professional society email networks will ideally capture that intended population.

### Description of survey tool and distribution

The online survey will contain a description of the study objectives, benefits, and harms of participating in the study, requirements for participation, the implications of the research findings, and a consent form. The survey will be generated using Qualtrics XM software (Provo, UT, USA) and administered to clinician emails via approved professional society mailing lists. There will be 30 questions requiring multiple choice responses with and without freeform text input. The survey will be divided into three sections: (1) demographics and surgical training, (2) the use of IOPTH and surgical adjuncts during parathyroidectomy, and (3) perceived barriers to adopting IOPTH monitoring (Appendix 1). There will be adaptive reasoning within the survey based upon respondents’ answers to certain questions. Neither patients nor the public were involved in the design of the survey. The survey was created with input from five content experts and was piloted for flow and clinical sensibility with five local surgeons, whereby further refinements were implemented [15]. Our piloting suggested that survey completion requires approximately 8 minutes. This survey study will be conducted and reported in accordance with the Checklist for Reporting Results of Internet E-Surveys (CHERRIES) [16].

Survey respondents will not be required to answer every survey question to complete and submit the survey. We estimate it will take 5–8 minutes to complete the entire survey. There will be no non-English translations of the survey as it will only be distributed within North America. All respondents will only be able to submit survey once. Survey completion will not be timed and there will be no incentives for completing the survey. We will not stratify survey distribution based upon demographic variables or training level. The survey will be distributed on a rolling basis over the course of 6 months and will be emailed at least twice during this interval. The study team will not have access to respondent emails, as this will be managed by the professional society. The planned study start date is March 28, 2024, with an anticipated completion date of August 31, 2024. Respondents will not receive their completed survey results but will be notified of publication through their respective professional society.

### Planned sample size

In Canada, there are an estimated 715 otolaryngology–head and neck surgeons practicing in either community or academic centres [17]. In the US, there are an estimated 12,609 otolaryngology– head and neck surgeons practicing in either community or academic centres [18]. There are 518 active or corresponding members in the AAES [14]. We do understand that not all practicing surgeons are members of the professional societies selected for survey distribution and thus, estimate that we will be sampling from a population of approximately 10,000 practicing surgeons and trainees across North America. As the goal of this survey is to describe IOPTH usage trends without a predefined primary outcome, we will not perform a sample size calculation. We will, however, aim for a survey completion rate of 60–75%, which is consistent with other survey studies of surgeons and will help maintain external validity [15, 19].

### Study outcomes

The overall goal of this survey study is to explore clinical indications and technical details for employing IOPTH monitoring in primary, secondary, and tertiary hyperparathyroidism. We plan on characterizing IOPTH usage trends in the context of type of surgical training, type of surgical practice, and geography of surgical practice. The respondent demographic questions will be explored using nominal variables, while indications and details regarding IOPTH monitoring will be explored using both nominal and ordinal variables (i.e., Likert scale responses).

### Data Analysis Plan

For all completed or partially completed surveys, we will perform descriptive analyses to report proportions and frequencies for each survey response. We will evaluate variability in responses and estimates using 95% confidence intervals. As all responses will be described as nominal or ordinal variables, we will perform chi-square or Fischer’s exact tests for comparative analysis with an emphasis on comparing type of surgical discipline and location of practice. Based on survey responses, we will also evaluate the relationship of surgical practice location and the year of acquisition on the uptake of IOPTH monitoring technology. We also plan to perform logistic regression analysis to determine the effect of demographic and training variables on responses pertaining to rationale and details for using IOPTH monitoring and barriers to obtaining IOPTH technology. The key characteristics that we will insert in our multiple logistic regression model will be age, gender, type/level of training, type of practice, surgical volume, and location of training and practice. We will utilize the stepwise regression analysis to include significant variables into the model. We will evaluate model assumptions and goodness-of-fit by examining residuals and at least 10 observations will be required for each independent variable to be included into the regression model. There will be no planned subgroup analyses. Any free-text responses will be evaluated based upon conceptual analysis. All statistical analyses will be performed using R (version 4.3.1).

## DISCUSSION

In 2016, the American Association of Endocrine Surgeons (AAES) strongly recommended the use of IOPTH in minimally invasive parathyroidectomy to reduce operative failure [1]. Furthermore, it has been suggested that IOPTH monitoring may be helpful in guiding challenging parathyroid surgery such as in cases of parathyroid hyperplasia and tertiary hyperparathyroidism [20, 21]. Outside of primary hyperparathyroidism, however, there is little standardization and consensus in the utility of IOPTH monitoring. The survey will explore alternative indications for applying IOPTH monitoring to guide parathyroidectomy and describe the technical details of using IOPTH monitoring at institutions across North America. These results will help to inform future trials aimed at optimizing IOPTH guidance in challenging parathyroid surgeries.

## ETHICS AND DISSEMINATION

### Ethical and Safety Considerations

Ethics approval was obtained from the Hamilton Integrated Research Ethics Board (HiREB) as project number 2023-17173-GRA. Participation in this study will be voluntary and all participants will provide informed consent before starting the online survey. Participation will unlikely cause any adverse effects or discomfort related to the conduct or outcomes of the survey. There will be no timer associated with survey completion. All data collected will be anonymous. Survey datasets will be securely stored in Qualtrics, protected by Single-Sign-On, and in password-protected OneDrive accounts. The data will reside on password-protected, encrypted personal computers accessed via secure networks. Data will be destroyed two years following the completion of the study.

### Protocol Amendments

All protocol amendments will be submitted to the HiREB as modifications prior to implementation. Amendments will also be communicated during dissemination to both academic and lay audiences.

### Dissemination Plan

The findings of the survey will be submitted for publication in a peer-reviewed journal aimed at informing surgical practices in head and neck, and endocrine surgery. The primary investigator will present findings at national and international conferences. We will also communicate study findings to relevant stakeholders (i.e., surgeons, universities, hospital administrations, and government officials) to brainstorm other research questions and translate survey results into actionable items. These results will be used to inform future observational studies, cost analyses, and trials aimed at improving the adoption and optimization of IOPTH technology for guiding parathyroidectomy.

## Data Availability

No datasets were generated or analysed during the current study. All relevant data from this study will be made available upon study completion

## APPENDICES

**Appendix 1**. Survey cover letter and questionnaire.

## Acknowledgements

None

## Contributors

PS, HZ, MA, MKG, and JP conceived the research question. The final version of this manuscript was approved by all authors. All authors have agreed to be held accountable for this work.

## Funding

None

## Competing interests

None

## Patient consent for publication

None

## Notes

Conflicts of interest: None

### Competing Interest Statement

The authors have declared no competing interest.

### Funding Statement

The author(s) received no specific funding for this work.

### Author Declarations

Hamilton integrated Research Ethics Board

## REFERENCES

1. Wilhelm SM, Wang TS, Ruan DT, Lee JA, Asa SL, Duh QY, et al. The American Association of Endocrine Surgeons Guidelines for Definitive Management of Primary Hyperparathyroidism. JAMA Surg. 2016;151(10):959–68. doi:10.1001/jamasurg.2016.2310. PubMed PMID: 27532368.

2. Bilezikian JP, Cusano NE, Khan AA, Liu JM, Marcocci C, Bandeira F. Primary hyperparathyroidism. Nat Rev Dis Primers. 2016;2:16033. Epub 2016/05/20. doi:10.1038/nrdp.2016.33. PubMed PMID: 27194212; PubMed Central PMCID: PMCPMC5385896.

3. Saliba W, El-Haddad B. Secondary hyperparathyroidism: pathophysiology and treatment. J Am Board Fam Med. 2009;22(5):574–81. doi:10.3122/jabfm.2009.05.090026. PubMed PMID: 19734404.

4. Wermers RA. Incidence of Primary Hyperparathyroidism in the Current Era: Have We Finally Reached a Steady State? J Clin Endocrinol Metab. 2023;108(12):e1749–e50. doi:10.1210/clinem/dgad267. PubMed PMID: 37170854; PubMed Central PMCID: PMCPMC10655524.

5. Torino C, Tripepi R, Russo D, Vilasi AD, Panuccio VA. Editorial: Secondary hyperparathyroidism: an ongoing challenge for the nephrologist. Front Med (Lausanne). 2023;10:1305791. Epub 20231123. doi:10.3389/fmed.2023.1305791. PubMed PMID: 38076250; PubMed Central PMCID: PMCPMC10704019.

6. Bozic M, Diaz-Tocados JM, Bermudez-Lopez M, Forne C, Martinez C, Fernandez E, Valdivielso JM. Independent effects of secondary hyperparathyroidism and hyperphosphataemia on chronic kidney disease progression and cardiovascular events: an analysis from the NEFRONA cohort. Nephrol Dial Transplant. 2022;37(4):663–72. doi:10.1093/ndt/gfab184. PubMed PMID: 34021359.

7. Steinl GK, Kuo JH. Surgical Management of Secondary Hyperparathyroidism. Kidney Int Rep. 2021;6(2):254–64. Epub 20201230. doi:10.1016/j.ekir.2020.11.023. PubMed PMID: 33615051; PubMed Central PMCID: PMCPMC7879113.

8. Ernst H, Sowerby L, Sahovaler A, Macneil D, Nichols A, Yoo J, et al. Rapid standardized operating rooms (RAPSTOR) in thyroid and parathyroid surgery. J Otolaryngol Head Neck Surg. 2021;50(1):44. Epub 20210708. doi:10.1186/s40463-021-00525-x. PubMed PMID: 34238389; PubMed Central PMCID: PMCPMC8265141.

9. Itani M, Middleton WD. Parathyroid Imaging. Radiol Clin North Am. 2020;58(6):1071–83. Epub 2020/10/13. doi:10.1016/j.rcl.2020.07.006. PubMed PMID: 33040849.

10. Irvin GL, 3rd, Dembrow VD, Prudhomme DL. Operative monitoring of parathyroid gland hyperfunction. Am J Surg. 1991;162(4):299–302. doi:10.1016/0002-9610(91)90135-z. PubMed PMID: 1683177.

11. Medas F, Cappellacci F, Canu GL, Noordzij JP, Erdas E, Calo PG. The role of Rapid Intraoperative Parathyroid Hormone (ioPTH) assay in determining outcome of parathyroidectomy in primary hyperparathyroidism: A systematic review and meta-analysis. Int J Surg. 2021;92:106042. Epub 20210730. doi:10.1016/j.ijsu.2021.106042. PubMed PMID: 34339883.

12. Quinn AJ, Ryan EJ, Garry S, James DL, Boland MR, Young O, et al. Use of Intraoperative Parathyroid Hormone in Minimally Invasive Parathyroidectomy for Primary Hyperparathyroidism: A Systematic Review and Meta-analysis. JAMA Otolaryngol Head Neck Surg. 2021;147(2):135–43. doi:10.1001/jamaoto.2020.4021. PubMed PMID: 33211086; PubMed Central PMCID: PMCPMC7677873.

13. Pitt SC, Sippel RS, Chen H. Secondary and tertiary hyperparathyroidism, state of the art surgical management. Surg Clin North Am. 2009;89(5):1227–39. doi:10.1016/j.suc.2009.06.011. PubMed PMID: 19836494; PubMed Central PMCID: PMCPMC2905047.

14. Beninato T, Laird AM, Graves CE, Drake FT, Alhefdhi A, Lee JA, et al. Impact of the COVID-19 pandemic on the practice of endocrine surgery. Am J Surg. 2022;223(4):670–5. Epub 20210721. doi:10.1016/j.amjsurg.2021.07.009. PubMed PMID: 34315576; PubMed Central PMCID: PMCPMC8294714.

15. Burns KE, Duffett M, Kho ME, Meade MO, Adhikari NK, Sinuff T, et al. A guide for the design and conduct of self-administered surveys of clinicians. CMAJ. 2008;179(3):245–52. doi:10.1503/cmaj.080372. PubMed PMID: 18663204; PubMed Central PMCID: PMCPMC2474876.

16. Eysenbach G. Improving the quality of Web surveys: the Checklist for Reporting Results of Internet E-Surveys (CHERRIES). J Med Internet Res. 2004;6(3):e34. Epub 20040929. doi:10.2196/jmir.6.3.e34. PubMed PMID: 15471760; PubMed Central PMCID: PMCPMC1550605.

17. Brandt MG, Scott GM, Doyle PC, Ballagh RH. Otolaryngology - Head and Neck Surgeon unemployment in Canada: a cross-sectional survey of graduating Otolaryngology - Head and Neck Surgery residents. J Otolaryngol Head Neck Surg. 2014;43(1):37. Epub 20140916. doi:10.1186/s40463-014-0037-3. PubMed PMID: 25683630; PubMed Central PMCID: PMCPMC4329541.

18. Hughes CA, McMenamin P, Mehta V, Pillsbury H, Kennedy D. Otolaryngology workforce analysis. Laryngoscope. 2016;126 Suppl 9:S5–S11. Epub 20160831. doi:10.1002/lary.26238. PubMed PMID: 27576957.

19. Meyer VM, Benjamens S, Moumni ME, Lange JFM, Pol RA. Global Overview of Response Rates in Patient and Health Care Professional Surveys in Surgery: A Systematic Review. Ann Surg. 2022;275(1):e75–e81. doi:10.1097/SLA.0000000000004078. PubMed PMID: 32649458; PubMed Central PMCID: PMCPMC8683255.

20. Thielmann A, Kerr P. Validation of selective use of intraoperative PTH monitoring in parathyroidectomy. J Otolaryngol Head Neck Surg. 2017;46(1):10. Epub 2017/02/09. doi:10.1186/s40463-017-0188-0. PubMed PMID: 28166819; PubMed Central PMCID: PMCPMC5294871.

21. Ahn D, Kwak JH. Role and Recent Trend of Intraoperative Parathyroid Hormone Monitoring During Parathyroidectomy in Patients With Primary Hyperparathyroidism. Korean Journal of Otorhinolaryngology-Head and Neck Surgery. 2022;65(5):253–9. doi:10.3342/kjorl-hns.2022.00332.

